# Early elevation of FIB-4 liver fibrosis score is associated with adverse outcomes among patients with COVID-19

**DOI:** 10.1101/2020.09.01.20186080

**Authors:** Fangfei Xiang, Jing Sun, Po-Hung Chen, Peijin Han, Haipeng Zheng, Shuijiang Cai, Gregory D. Kirk

## Abstract

**Background:** Limited prior data suggest that pre-existing liver disease was associated with adverse outcomes among patients with COVID-19. FIB-4 is a noninvasive index of readily available laboratory measurements that represents hepatic fibrosis. The association of FIB-4 with COVID-19 outcomes has not been previously evaluated.

**Methods:** FIB-4 was evaluated at admission in a cohort of 267 patients admitted with early-stage COVID-19 confirmed through RT-PCR. Hazard of ventilator use and of high-flow oxygen was estimated using Cox regression models controlled for covariates. Risk of progress to severe cases and of death/prolonged hospitalization (>30 days) were estimated using logistic regression models controlled for same covariates.

**Results:** Forty-one (15%) patients progressed to severe cases, 36 (14%) required high-flow oxygen support, 10 (4%) required mechanical ventilator support, and 1 died. Patients with high FIB-4 score (>3.25) were more likely to be older with pre-existing conditions. FIB-4 between 1.45-3.25 was associated with over 5-fold (95% CI: 1.2-28) increased hazard of high-flow oxygen use, over 4-fold (95% CI: 1.5-14.6) increased odds of progress to severe stage, and over 3-fold (95% CI: 1.4-7.7) increased odds of death or prolonged hospitalization. FIB-4>3.25 was associated with over 12-fold (95% CI: 2.3-68. 7) increased hazard of high-flow oxygen use and over 11-fold (95% CI: 3.1-45) increased risk of progress to severe disease. All associations were independent of sex, number of comorbidities, and inflammatory markers (D-dimer, C-reactive protein).

**Conclusions:** FIB-4 at early-stage of COVID-19 disease had an independent and dose-dependent association with adverse outcomes during hospitalization. FIB-4 provided significant prognostic value to adverse outcomes among COVID-19 patients.

## INTRODUCTION

Patients infected with Severe Acute Respiratory Syndrome coronavirus-2 (SARS-CoV-2), the virus causing Coronavirus Disease 2019 (COVID-19), present a wide spectrum of disease severity, from asymptomatic to mild to critically ill requiring organ support and associated with high mortality [1, 2]. Early predictors of disease progression are critical for COVID-19 disease management; identifying high risk individuals can inform appropriate triage and allocate limited healthcare resources appropriately during this global pandemic. Previous studies identified that patients with multi-morbidities and of older age had worse prognosis [1, 3-5]. However, the role of liver disease in COVID-19 remains unclear.

Recent studies have reported that liver enzyme abnormalities and acute liver injury were common among patients with severe COVID-19 infection [6-10]. Liver dysfunction is likely multifactorial, and could result from liver injury directly by the SARS-CoV-2 virus, by the associated cytokine storm, by resulting multi-organ failure, or by medications used to manage patients with COVID-19 [7, 11]. Acute liver decompensation following COVID-19 infection could also represent exacerbation of pre-existing chronic liver diseases [12]. Liver dysfunction and cirrhosis are associated with innate immune dysfunction which could enhance susceptibility to acute proinflammatory responses [13]. These pathways are hypothesized to lead to severe outcomes among patients with COVID-19.

Current guidelines recommend prioritizing testing for COVID-19 and early admission for patients with chronic liver disease [12]. However, there remain limited data explaining whether individuals with pre-existing liver disease might have elevated risk of severe symptoms and worse clinical outcomes with COVID-19 [7, 11, 12, 14-16]. Routine clinical encounters often do not include detailed imaging evaluation of liver function, which increases the challenge of identifying pre-existing liver condition in early COVID-19 disease stage. Further, while liver injury is a prominent feature of severe COVID-19, the impact of liver injury at early-stage COVID-19 remains largely unexplored.

The Fibrosis-4 (FIB-4) index was developed to leverage inexpensive and routinely available laboratory results (platelet count, aspartate transaminase [AST], and alanine transaminase [ALT]) along with age to noninvasively identify liver fibrosis among patients with viral hepatitis [17-19]. FIB-4 has demonstrated a high predictive value for multiple liver-related outcomes (hepatocellular carcinoma, severe fibrosis, cirrhosis verified by biopsy) in patients with viral hepatitis [20, 21]. FIB-4 also predicts all-cause inpatient mortality and all liver-related outcomes among individuals without known chronic liver diagnosis [22, 23]. In the absence of viral hepatitis infection, a previous study also showed an association between FIB-4 and HIV infection, suggesting a potential relationship between HIV infection and hepatic fibrosis [24]. We sought to evaluate the association of FIB-4 scores at an early-stage of SARS-CoV-2 infection with adverse outcomes during hospitalization.

## METHODS

### Study Participants and Case Definition

We conducted a retrospective cohort study of adults with confirmed SARS-CoV-2 infection admitted to a leading hospital specializing in infectious diseases (Guangzhou No. 8 People’s Hospital) in the capital of Guangdong province, China from January 20 to February 10, 2020. During this time period, suspected cases of COVID-19 in Guangzhou city were evaluated at the hospital and admitted immediately after confirmed diagnosis, regardless of the presence of symptoms. Suspected cases were identified through (1) symptom presentation, (2) contact tracing of confirmed cases, (3) testing individuals with a recent travel history to Wuhan, China whom were under mandatory quarantine. All cases included in this analysis were confirmed by RT-PCR (Daan Gene Co., Sun Yat-sen University) of nasopharyngeal swabs. A total of 274 patients were admitted during the recruitment period. As our focus was on early-stage disease, we excluded 7 patients with respiratory decompensation (room air oxygen saturation <93% or respiratory rate ≥30 breaths per minute) at admission. Compared to the 267 persons included in the analysis, the 7 patients excluded with respiratory decompensation at admission were more likely to be older, diabetic, and have liver disease (all p<0.05; data not shown). This study was approved by the Institutional Review Board of Guangzhou No. 8 People’s Hospital.

### Outcomes

Outcomes, exposures of interest, and other covariates were identified through standardized abstraction from medical records by three trained physicians. Four primary adverse outcomes were assessed in four different statistical models: (1) time to use of high-flow oxygen, (2) time to ventilator use, (3) risk of severe disease stage at any time during hospitalization, and (4) risk of either death or prolonged hospitalization (>30 days). Severe COVID-19 disease was defined by ≥1 of the following occurring at any point during hospitalization: 1) respiration rate ≥30 breaths/minute; 2) resting oxygen saturation <93%; 3) ratio of arterial oxygen partial pressure (PaO_2_ in mmHg) to fractional inspired oxygen (FiO_2_, expressed as fraction) ≤300mmHg; or 4) imaging demonstrating rapid lung disease progression within 48 hours.

### FIB-4 Measurement at Baseline

FIB-4 score was calculated using age, AST (U/L), ALT (U/L), and platelet count (10^9^/*L*) using standard methods [17-19]. All laboratory values were obtained within 24 hours of admission to the hospital. FIB-4 scores were categorized according to prior cutpoints validated to be a proxy of Ishak stage of normal (0-1), significant (2-3), and advanced (4-6) hepatic fibrosis (<1.45, 1.45-3.25, and >3.25, respectively) [17-19].

### Other Covariates of Interest

Symptoms and signs on admission were obtained through self-report (diarrhea, fatigue, coughing, mucus) or physical examination (fever, rales). Self-report of pre-existing conditions including high blood pressure, diabetes, heart disease, liver disease, kidney disease, psychological disorders, and other major diseases (including neurological disorders, cancers, other gastrointestinal tract diseases, and metabolic diseases; N=26) were obtained by physicians during the admission encounter. Pre-existing comorbidities were categorized into: none, 1, and ≥2 conditions. Complete blood count (red blood cell, white blood cell, neutrophil, lymphocyte, monocyte, hemoglobin, and platelet counts), inflammatory markers (D-dimer and C-reactive protein), liver panel (ALT, AST, total bilirubin, direct bilirubin, albumin, and prothrombin time), kidney panel (creatinine, urine albumin, glomerular filtration rate [GFR], and cystatin C), and other enzymes (lactate dehydrogenase, creatine kinase, and creatine kinase myocardial band [MB]) were routinely measured upon admission. Values that had substantial missing data (e.g., SS-A/Ro antibodies) were excluded from the analysis; all laboratory tests included in the analysis had ≤ 10% missing. Elevated ALT and AST were defined using thresholds based on American College of Gastroenterology guidelines [25].

### Statistical Analysis

Statistical differences in baseline characteristics and biochemical panels by FIB-4 score groups (<1.45, 1.45-3.25, >3.25) were compared using two-sided T test, Kruskal-Wallis equality-of-populations rank test, or Chi-square test. Horizontal line plots were used to visualize disease trajectories of all patients by FIB-4 categories [26, 27]. Cumulative hazard curves with 95% confidence interval (95% CI) were used to illustrate time from admission to ventilator use. Cumulative hazard functions by FIB-4 groups were tested by log-rank test for trend. Time-to-event analysis by FIB-4 groups was estimated using Cox regression models for two endpoints: [a] ventilator use or [b] high-flow oxygen use, adjusting for gender, number of comorbidities, time from symptom onset to admission, and inflammatory markers. Person-time at risk for adverse outcomes (ventilator or high-flow oxygen) was calculated from the date of admission to the date of event, discharge (among those without adverse events), or March 31st, 2020 (among individuals still in care). Risk of severe outcomes by FIB-4 groups was evaluated using multivariable logistic regression models with two end-points: [a] progression to severe disease stage (as defined above based on objective clinical and imaging criteria) at any point during hospitalization vs. only had mild or no symptoms during hospitalization; [b] death or prolonged hospitalization vs. discharged within 30 days. Models were adjusted for gender, number of comorbidities, time from symptom onset to admission, and inflammatory markers. As age is included in the FIB-4 calculation, we did not further adjust models for age to avoid collinearity. Statistical analysis and figures were performed in STATA 15 (College Station, TX) and R.

## RESULTS

### Patient characteristics

The patient population studied included individuals tested because of potential exposure (124, 45.3%) or because of symptoms (150, 54.7%). In this setting in China in the early-stage of the pandemic, all persons with a positive SARS-CoV-2 test were admitted for observation; 17% of the patients had no symptoms at admission. The median time from symptom onset to admission was ≤4 days (interquartile range, IQR: 0 – 7 days). **Figure 1** illustrates the disease trajectory of each individual patient. Of 267 patients, 41 (15.4%) progressed to severe disease stage, 36 (13.5%) required high-flow oxygen support, 10 (3.7%) required mechanical ventilator support, and 7 (2.6%) required extracorporeal membrane oxygenation (ECMO) support during hospitalization. By March 31^st^, 2020, 1 patient had died, 5 were still in care, and the remainder had been discharged.

**Figure 1.**
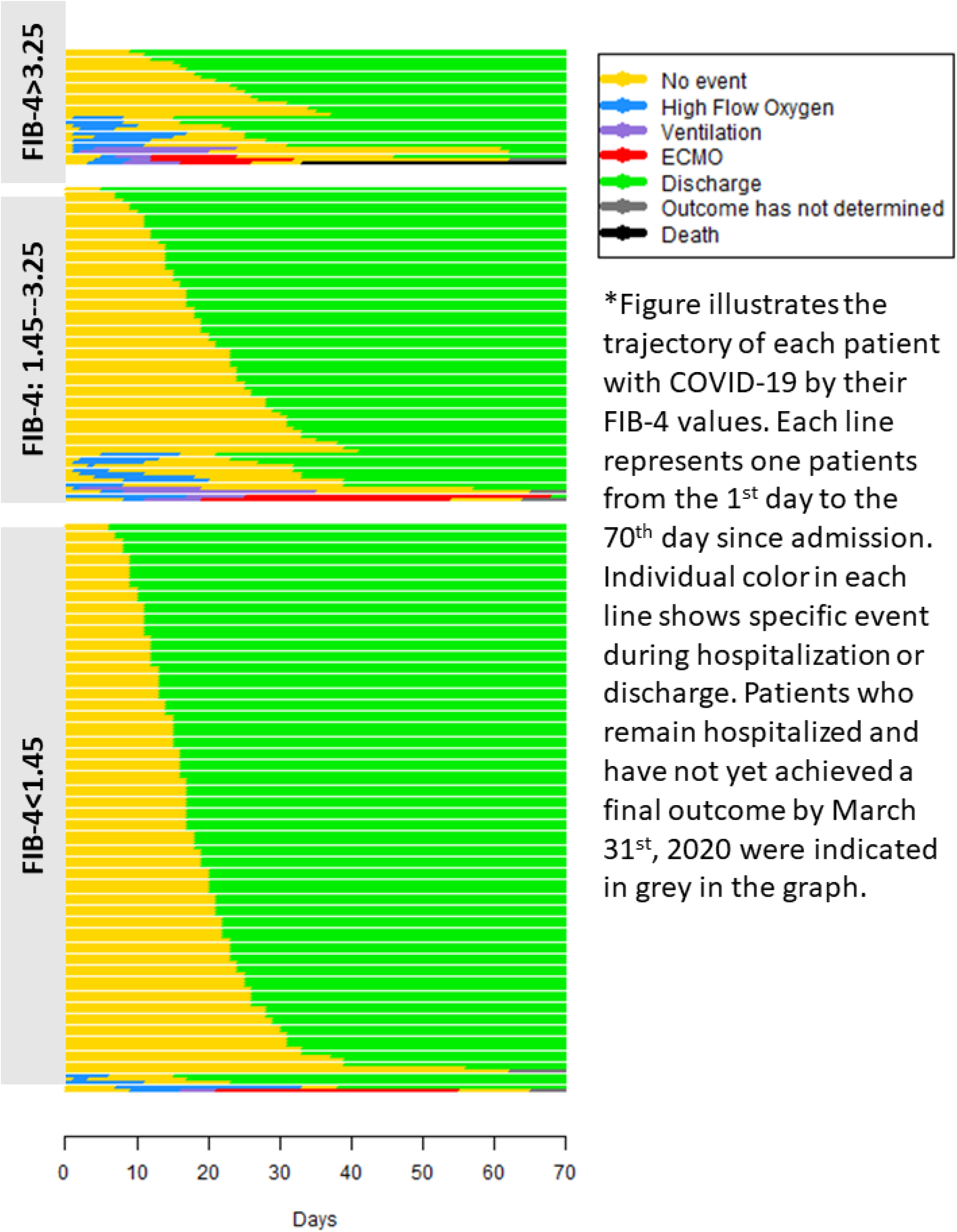
Disease trajectories among patients with COVID-19 by FIB-4 groups at admission.

Among all patients, 153 (57.3%) had FIB-4 <1.45, 89 (33.3%) had values between 1.45-3.25, and 32 (12.0%) had FIB-4 >3.25. Individuals who were older, had heavy alcohol consumption, or had a greater number of pre-existing conditions were more likely to have higher baseline FIB-4 scores (**Table 1**, all P<0.05). In terms of clinical presentation, patients with higher FIB-4 scores were more likely to have fever and fatigue at admission; other symptoms were not related to FIB-4 scores.

**Table 1.** Characteristics of patients at time of admission overall and by FIB-4 score category.

|  | Total<br>N=267 | FIB-4 score |  |  | p-value |
| --- | --- | --- | --- | --- | --- |
|  |  | <1.45<br>N=153 (57.3%) | 1.45-3.25<br>N=89 (33.3%) | >3.25<br>N=32 (12.0%) |  |
| **Demographic and health behavior** |  |  |  |  |  |
| Age, mean (SD) | 47.7 (16.9) | 39.1 (14.6) | 57.0 (11.6) | 65.1 (12.2) | <0.001 |
| Female sex, N (%) | 145 (54.5%) | 82 (54.3%) | 48 (57.1%) | 15 (48.4%) | 0.70 |
| Heavy smoker (10 cigs/day)<sup>a</sup>, N (%) | 6 (2.2%) | 0 (0.0%) | 5 (6.0%) | 1 (3.2%) | 0.012 |
| Heavy alcohol consumption<sup>a</sup>, N (%) | 2 (0.7%) | 0 (0.0%) | 2 (2.4%) | 0 (0.0%) | 0.11 |
| **Baseline pre-existing conditions** |  |  |  |  |  |
| Hypertension, N (%) | 50 (18.7%) | 18 (11.8%) | 24 (28.6%) | 8 (25.8%) | 0.004 |
| Diabetes, N (%) | 15 (5.6%) | 4 (2.6%) | 9 (10.7%) | 2 (6.5%) | 0.035 |
| Heart disease, N (%) | 11 (4.1%) | 1 (0.7%) | 5 (6.0%) | 5 (16.1%) | <0.001 |
| Liver disease, N (%) | 6 (2.2%) | 1 (0.7%) | 1 (1.2%) | 4 (12.9%) | <0.001 |
| Kidney disease, N (%) | 4 (1.5%) | 1 (0.7%) | 1 (1.2%) | 2 (6.5%) | 0.05 |
| Psychological disorder, N (%) | 1 (0.4%) | 1 (0.7%) | 0 (0.0%) | 0 (0.0%) | 0.68 |
| Other diseases, N (%) | 25 (9.4%) | 7 (4.6%) | 13 (15.5%) | 5 (16.1%) | 0.009 |
| No. of Comorbidities, N (%) |  |  |  |  | <0.001 |
| none | 183 (68.5%) | 125 (82.2%) | 45 (53.6%) | 13 (41.9%) |  |
| 1 | 61 (22.8%) | 21 (13.8%) | 28 (33.3%) | 12 (38.7%) |  |
| 2 or more | 23 (8.6%) | 6 (3.9%) | 11 (13.1%) | 6 (19.4%) |  |
| **Symptoms at admission** |  |  |  |  |  |
| Fever, N (%) | 186 (69.7%) | 94 (61.8%) | 65 (77.4%) | 27 (87.1%) | 0.004 |
| Coughing, N (%) | 151 (56.6%) | 81 (53.3%) | 47 (56.0%) | 23 (74.2%) | 0.10 |
| Mucus, N (%) | 61 (22.9%) | 28 (18.5%) | 23 (27.4%) | 10 (32.3%) | 0.13 |
| Rales, N (%) | 0 (0.0%) | 0 (0.0%) | 0 (0.0%) | 0 (0.0%) | - |
| Diarrhea, N (%) | 6 (2.2%) | 2 (1.3%) | 3 (3.6%) | 1 (3.2%) | 0.50 |
| Fatigue, N (%) | 17 (6.4%) | 2 (1.3%) | 9 (10.7%) | 6 (19.4%) | <0.001 |

\***Boldface** indicates statistically significant difference by baseline FIB-4 score categories previously validated to be indicative of none, significant and advanced fibrosis.
<sup>a</sup>Heavy smoker was defined as consuming 10 cigarettes per day; heavy alcohol consumption was defined as 20g/day for female and 40g/day for male for at least 5 years.

### Hematologic and biochemical parameters and FIB-4 score

The majority of ALT and AST levels were within the normal range at admission (median, [IQR]: 20.3 [14-30], 20.3 [16.5-28.9], respectively; **Table 2**). In addition to higher AST and lower platelet counts that are incorporated into the FIB-4 index, patients with higher FIB-4 scores were more likely to have higher direct bilirubin and lower albumin; ALT or prothrombin time were not different by FIB-4 score. Several biochemical and hematologic abnormalities differed by FIB-4 scores in a dose-dependent manner, notably including reduced total lymphocyte count and hemoglobin levels and increased C-reactive protein (CRP), lactic acid dehydrogenase, creatinine kinase and serum cystatin C (**Table 2**).

**Table 2.** Hematologic and biochemical parameters at admission by FIB-4 score category.

|  |  | FIB-4 categories |  |  |  |
| --- | --- | --- | --- | --- | --- |
|  | Overall | <1.45 | 1.45-3.25 | >3.25 | p-value<sup>a</sup> |
| RBC, million/mm<sup>3</sup>, mean (SD) | 4.6 (2.3) | 4.9 (2.9) | 4.4 (0.6) | 4.2 (0.6) | 0.13 |
| WBC, k/mm<sup>3</sup>, mean (SD) | 5.0 (1.8) | 5.2 (1.6) | 4.7 (2.0) | 5.0 (1.7) | 0.16 |
| Neutrophils, k/mm<sup>3</sup>, mean (SD) | 3.0 (1.5) | 3.0 (1.3) | 3.0 (1.7) | 3.4 (1.7) | 0.27 |
| Lymphocyte, k/mm<sup>3</sup>, mean (SD) | **1.5 (0.7)** | **1.7 (0.8)** | **1.3 (0.5)** | **1.2 (0.6)** | **<0.001** |
| Monocyte, k/mm<sup>3</sup>, mean (SD) | 0.4 (0.2) | 0.4 (0.2) | 0.4 (0.2) | 0.3 (0.1) | 0.25 |
| Hemoglobin, g/dL, mean (SD) | **13.6 (1.7)** | **13.8 (1.6)** | **13.5 (1.7)** | **12.9 (2.1)** | **0.023** |
| Platelet count, k/mm<sup>3</sup>, mean (SD) | **189.2 (65.0)** | **219.1 (60.3)** | **153.2 (42.7)** | **140.6 (59.7)** | **<0.001** |
| D-dimer, µg/mL, mean (SD) | 1.1 (4.7) | 0.8 (1.6) | 1.7 (7.9) | 0.9 (0.6) | 0.38 |
| CRP*, mean (SD) | **2.8 (0.7)** | **2.6 (0.5)** | **2.9 (0.8)** | **3.5 (0.8)** | **<0.001** |
| ***Liver panel*** |  |  |  |  |  |
| ALT |  |  |  |  |  |
| Original, U/L, median (IQR) | 20.3 (14-30) | 20 (13.4-27.1) | 19.4 (14.3-40.4) | 25 (16.2-42.8) | 0.20<sup>b</sup> |
| Log transferred*, mean (SD) | 3.1 (0.6) | 3.0 (0.5) | 3.1 (0.7) | 3.2 (0.7) | 0.11 |
| AST |  |  |  |  |  |
| Original, U/L, median (IQR) | **20.3 (16.5-28.9)** | **17.6 (14.9-22)** | **23.5 (18.7-35.1)** | **46 (28-58)** | **<0.001<sup>b</sup>** |
| Log transferred*, mean (SD) | **3.1 (0.5)** | **2.9 (0.4)** | **3.3 (0.5)** | **3.8 (0.6)** | **<0.001** |
| Total bilirubin, µmol/L, mean (SD) | 10.5 (6.4) | 10.0 (5.7) | 11.4 (7.6) | 10.6 (7.1) | 0.35 |
| Direct bilirubin, µmol/L, mean (SD) | **4.4 (2.6)** | **4.0 (1.9)** | **5.2 (3.6)** | **4.2 (1.6)** | **0.015** |
| Albumin, g/dL, mean (SD) | **39.7 (5.8)** | **41.5 (5.4)** | **38.0 (4.3)** | **33.0 (7.1)** | **<0.001** |
| Prothrombin time, second, mean (SD) | 13.5 (0.9) | 13.5 (0.8) | 13.4 (0.9) | 13.5 (1.3) | 0.45 |
| ***Kidney function*** |  |  |  |  |  |
| Creatinine, mmol/L, mean (SD) | 64.2 (26.8) | 61.6 (19.3) | 67.1 (26.5) | 69.5 (52.0) | 0.18 |
| Urine Albumin*, mean (SD) | **5.7 (0.4)** | **5.7 (0.3)** | **5.6 (0.4)** | **5.6 (0.5)** | **0.039** |
| GFR, mL/min/1.73 m<sup>2</sup>, mean (SD) | **155.3 (103.3)** | **168.8 (128.4)** | **133.3 (41.7)** | **151.8 (72.7)** | **0.044** |
| Cystatin C, mg/L, mean (SD) | **1.0 (0.7)** | **0.8 (0.2)** | **1.1 (1.1)** | **1.2 (0.7)** | **0.002** |
| ***Biochemical panel*** |  |  |  |  |  |
| Lactate dehydrogenase, U/L, mean (SD) | **210.3 (90.8)** | **187.1 (64.9)** | **225.3 (85.9)** | **276.7 (149.5)** | **<0.001** |
| Creatine kinase, U/L, mean (SD) | **118.1 (249.1)** | **86.1 (57.9)** | **113.4 (116.0)** | **276.7 (664.4)** | **<0.001** |
| Creatine kinase MB, IU/L, mean (SD) | 11.5 (7.9) | 11.9 (6.0) | 11.2 (6.6) | 10.4 (15.7) | 0.61 |

\*Values have been log transferred.
<sup>a</sup>P-value tested by two-sided T test unless indicated otherwise.<sup>b</sup>P-value tested by Kruskal-Wallis equality-of-populations rank test.

### FIB-4 and adverse outcomes

As demonstrated in **Figure 1**, despite fewer patients in the higher FIB-4 groups, more adverse clinical events and longer hospitalization were observed (median days of hospitalization [IQR] of 17 [12.5-23]; 21 [14-29.5]; and 25 [17-33], respectively among FIB-4<1.45; 1.45-3.25; and >3.25). Higher FIB-4 scores at admission were associated with increased hazard of ventilator use, high-flow oxygen, increased risk of progression to severe disease stage, and to death or prolonged hospitalization (**Table 3**). In particular, compared to FIB-4<1.45, FIB-4>3.25 was associated with over 23-fold (95% CI: 2.5-219) increased hazard of ventilator use, a 25-fold (95% CI: 6.8-98) increased hazard of using high-flow oxygen, ~21-fold (95% CI: 6.9-60.8) increased risk of progress to severe cases, and over 5-fold (95% CI: 1.9-13.2) increased odds of death or prolonged hospitalization. All associations were independent from sex, number of total comorbidities at admission, and time from symptoms onset to admission. After controlling for inflammatory markers (D-dimer, CRP), the associations between FIB-4 and adverse outcomes were attenuated but not eliminated (**Table 3**). Similar associations were observed among patients with FIB-4 between 1.45-3.25 compared to those with FIB-4 <1.45, although the magnitude of association was lower compared to patients with FIB-4 >3.25 (**Table 3**). Further demonstrations in **Figure 2** showed patients with higher FIB-4 were associated with higher hazard of using ventilators (p-value of test for trend of cumulative hazard functions<0.001). These results suggested a strong, dose-dependent association between high FIB-4 score at an early-stage of infection and adverse clinical outcomes among patients with COVID-19, and this association was not completely driven by acute inflammation.

**Table 3.**
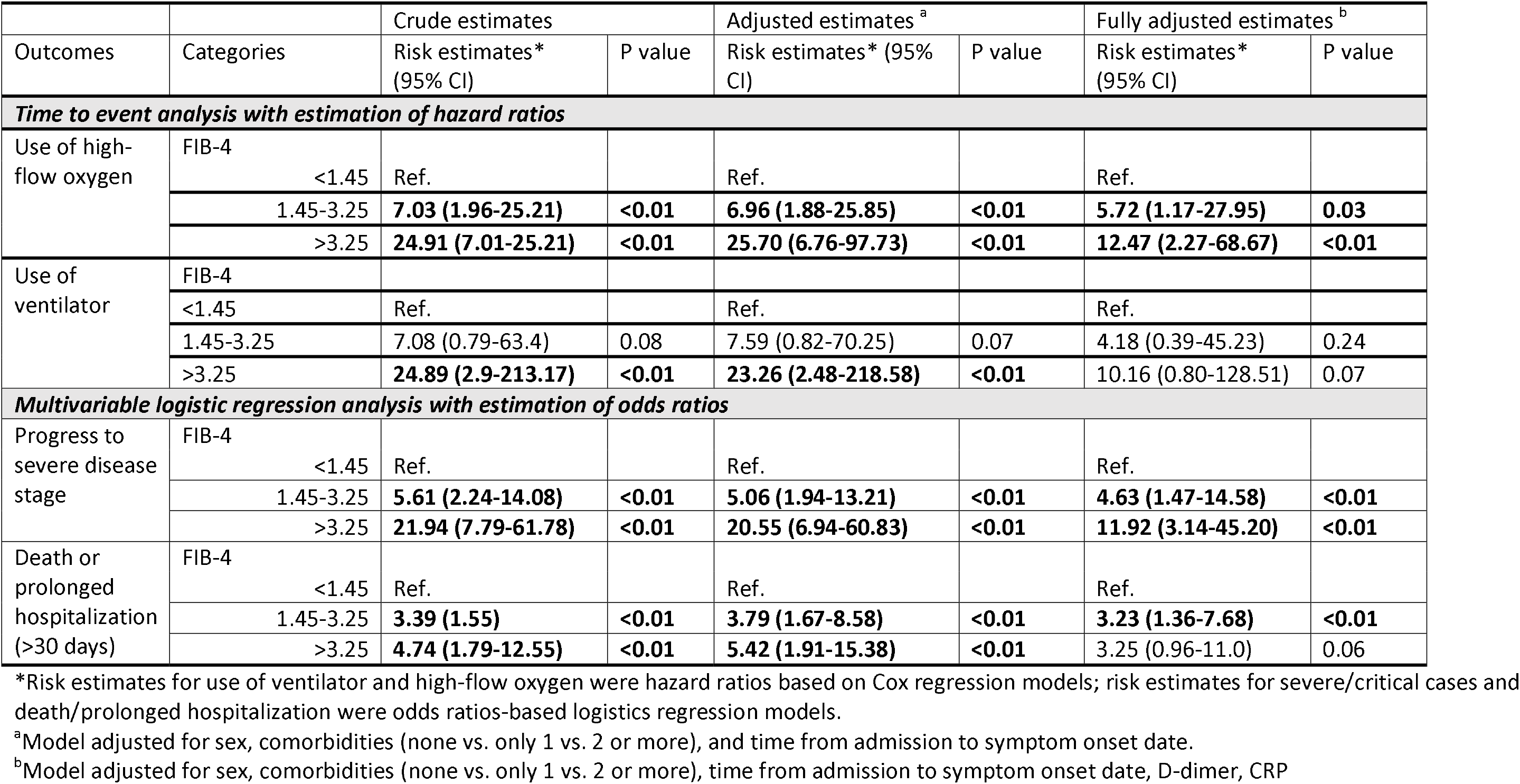
FIB-4 score category at admission and subsequent adverse outcomes during hospitalization among COVID-19 patients

**Figure 2.**
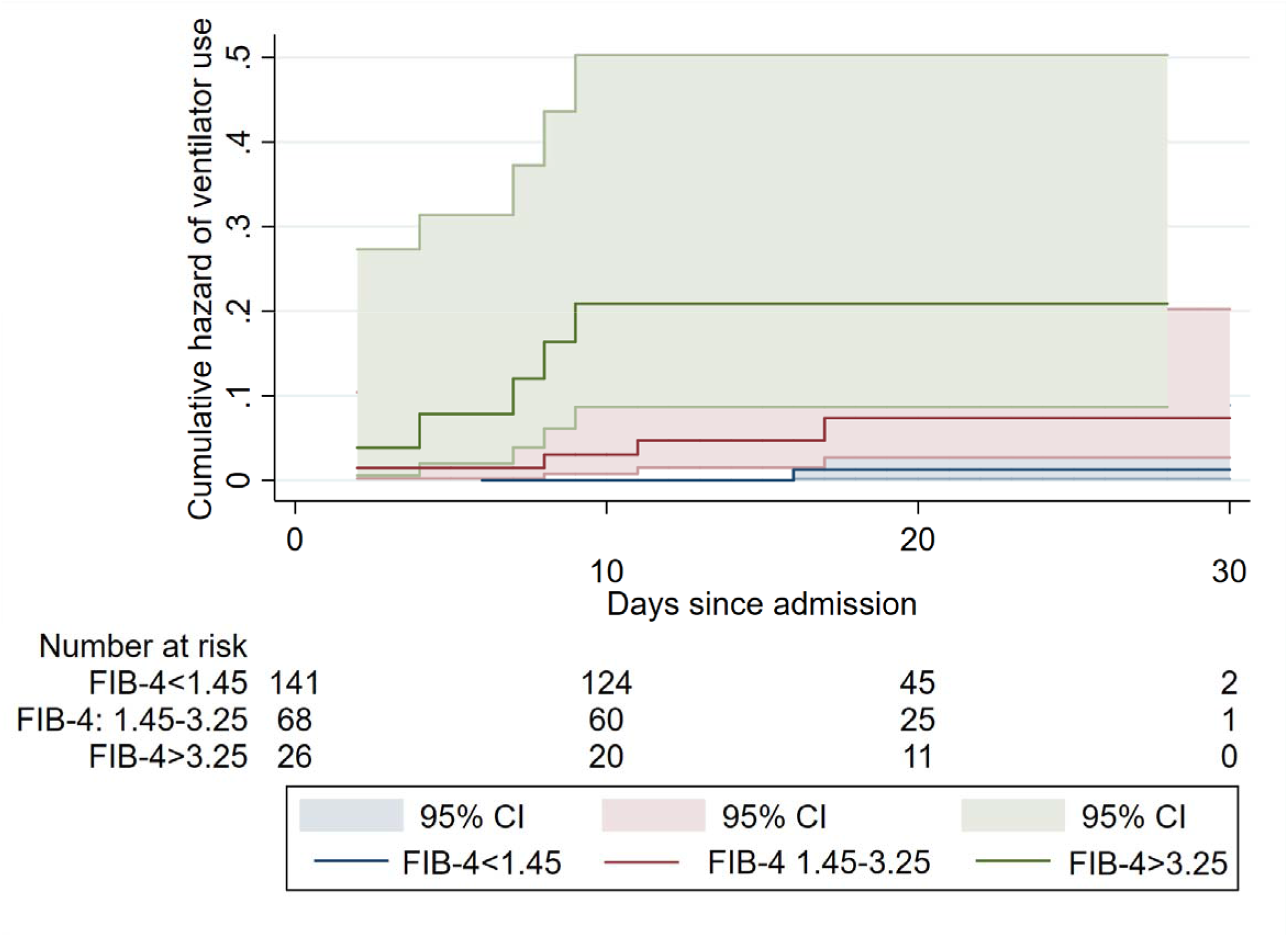
Cumulative hazard and 95% confidence interval (95% Cl) of ventilator requirement within 30 days since admission by FIB-4 score.

### Self-report liver disease, liver enzyme abnormalities and adverse outcomes

Self-reported liver disease (N=6) was associated with higher probability of death or prolonged hospitalization (**Supplementary Table S-1**). The lack of association in models on other adverse outcomes might suggest under-reporting of liver disease and lack of sufficient statistical power to observe the association. Abnormalities of ALT and AST alone were not independently associated with adverse outcomes during hospitalization. While elevated AST (>40 U/L) was associated with hazard of high-flow oxygen use and progress to severe cases during hospitalization, the association was attenuated after controlling for D-dimer and CRP, suggesting the association between single value elevation and adverse outcomes might be driven by acute inflammation (**Supplementary Table S-1**).

## DISCUSSION

We observed that FIB-4 scores at an early-stage of COVID-19 infection were independently and dose-dependently associated with multiple adverse outcomes during hospitalization. These results suggested that baseline hepatic fibrosis may be an early predictor of adverse outcomes among patients with COVID-19, and patients with chronic liver disease may have greater susceptibility to progressive disease.

Although there were only 6 patients who self-reported to have had a liver diagnosis prior to their COVID-19 infection, a substantial burden of liver disease related to chronic hepatitis B infection has been observed in the Chinese population in previous studies [28, 29]. Chronic infections of hepatitis B were observed in 8-12% of Chinese adults age 20 years and older [28], and infection rates may be much higher among older populations [29]. It is highly likely that the self-reported liver disease in the current study underestimated the true prevalence of underlying liver disease within this group. That might explain the high proportion of individuals with FIB-4>1.45 within this population.

Previous studies suggested pre-existing liver diseases (cirrhosis and NAFLD) were associated with higher mortality and faster disease progression among patients with COVID-19 [30-32] and other acute respiratory distress syndromes (ARDS) [33]. Several potential pathways could explain the association between underlying liver disease and adverse outcomes among people with COVID-19 [34-36]. Severe liver diseases were associated with innate immune dysfunction and could lead to enhanced susceptibility to an acute proinflammatory response [13]. These pathways can promote alveolar epithelial injury and increase vascular permeability among patients with ARDS and induce local and systemic inflammation, which eventually could lead to adverse clinical outcomes [34-36]. Additionally, patients with cirrhosis have high risk of multi-organ failure and increased mortality during infection [37], which may explain the increased likelihood of death or prolonged hospitalization in the current study. These findings underscored the importance of identifying underlying liver conditions among patients with COVID-19 in early-stage.

The current study has several limitations. We were not able to confirm liver fibrosis using ultrasound, elastography, or biopsy to better characterize liver disease severity among patients. Therefore, the association should be interpreted with caution. In addition, this is a single-site study with relatively low sample size. Thus, the external validity should be further evaluated in other cohorts with larger sample sizes. However, despite the limited sample size, we were still able to observe strong associations between high FIB-4 scores early on and multiple adverse outcomes during follow-up. Third, FIB-4 score comprised platelet count, ALT, and AST, which could fluctuate especially during acute inflammation, and severe COVID-19 cases could have elevated aminotransferases and thrombocytopenia during infection [1, 11]. Elevated FIB-4 score could be a product of acute inflammation rather than a reflection of hepatic fibrosis. However, we have limited our study cohort to only patients in the early-stage of COVID-19 infection, and the distributions of ALT and AST at admission were within the normal range in our study. We also controlled for multiple inflammatory markers in our final models. Based on our results, the independent association between FIB-4 score and adverse outcomes was not confounded or completely mediated by the inflammatory markers at admission, so the inflammation alone is unlikely to explain this association. Furthermore, even if FIB-4 is not fully representative of the underlying liver condition before the diagnosis of COVID-19, using FIB-4 at an early-stage of infection to predict adverse outcomes is still more sensitive compared to other single laboratory values (e.g., ALT, AST).

There are also several strengths to our study. By limiting the study to those patients who were still in the early-stage of infection and assessing the FIB-4 score at admission, the impact of drug-induced hepatic toxicities has been eliminated and lead-time bias has been minimized in the current study. Most studies on liver disease and COVID-19 were focused on liver injury during COVID-19 infection, and many were conducted among patients with progressive disease [9, 10]. In contrast, our cohort admitted all individuals as soon as they received a positive result of COVID-19, regardless of the presence of symptoms, which provided a unique opportunity for us to evaluate the natural history of disease progression based on early symptoms and characteristics. Lastly, although this is a single-site study, we were able to include all patients with COVID-19 in a large metropolitan city identified through strict contact tracing and during a period of mandatory quarantine, which prevented sampling bias.

In conclusion, FIB-4 is a noninvasive, inexpensive, and sensitive marker to predict disease progression among patients with COVID-19. Given FIB-4 scores are easily available in most clinical settings, wide screening of liver fibrosis using FIB-4 among patients with COVID-19 may provide important insights to define high-risk groups and improve clinical outcomes. Underlying liver disease may place individuals with COVID-19 at greater risk to develop adverse outcomes. Therefore, healthcare providers should emphasize self-quarantine and preventive measures to patients with severe liver fibrosis. Further research using a larger sample size is warranted to better understand the mechanisms of underlying liver condition and poor prognosis among patients with COVID-19.

## Data Availability

Data is available upon requested. Please contact Dr. Jing Sun for additional detail.

## Author contributions

**Study concept, study design:** JS, FX

**Manuscript writing:** JS

**Data acquisition, data collection:** FX, HZ, SC

**Data analysis and data management:** JS, PJH

**Critical review and edits:** GDK, PHC

## REFERENCES

1. Guan, W.-j., et al., Clinical characteristics of coronavirus disease 2019 in China. New England journal of medicine, 2020. 382(18): p. 1708–1720.

2. Long, Q.-X., et al., Clinical and immunological assessment of asymptomatic SARS-CoV-2 infections. Nature Medicine, 2020: p. 1–5.

3. Wang, B., et al., Does comorbidity increase the risk of patients with COVID-19: evidence from meta-analysis. Aging (Albany NY), 2020. 12(7): p. 6049.

4. Huang, C., et al., Clinical features of patients infected with 2019 novel coronavirus in Wuhan, China. The lancet, 2020. 395(10223): p. 497–506.

5. Ward, J.W. and M. Carlos del Rio, The COVID-19 Pandemic: An Epidemiologic, Public Health, and Clinical Brief. Clinical Liver Disease, 2020. 15(5): p. 170–174.

6. Fan, Z., et al., Clinical features of COVID-19-related liver damage. Clinical Gastroenterology and Hepatology, 2020.

7. Zhang, C., L. Shi, and F.-S. Wang, Liver injury in COVID-19: management and challenges. The Lancet Gastroenterology & Hepatology, 2020. 5(5): p. 428–430.

8. Cai, Q., et al., COVID-19: Abnormal liver function tests. Journal of hepatology, 2020.

9. Lei, F., et al., Longitudinal association between markers of liver injury and mortality in COVID - 19 in China. Hepatology, 2020.

10. Phipps, M.M., et al., Acute Liver Injury in COVID-19: Prevalence and Association with Clinical Outcomes in a Large US Cohort. Hepatology, 2020.

11. Bangash, M.N., J. Patel, and D. Parekh, COVID-19 and the liver: little cause for concern. The Lancet Gastroenterology & Hepatology, 2020.

12. Lau, G. and J.W. Ward, Synthesis of Liver Associations Recommendations for Hepatology and Liver Transplant Care During the COVID-19 Pandemic. Clinical Liver Disease, 2020. 15(5): p. 204–209.

13. Noor, M.T. and P. Manoria, Immune dysfunction in cirrhosis. Journal of clinical and translational hepatology, 2017. 5(1): p. 50.

14. Garrido, I., R. Liberal, and G. Macedo, “Review article: COVID-19 and liver disease-what we know on 1st May 2020”. Alimentary Pharmacology & Therapeutics, 2020.

15. Tian, S., et al., Pathological study of the 2019 novel coronavirus disease (COVID-19) through postmortem core biopsies. Modern Pathology, 2020: p. 1–8.

16. Boettler, T., et al., Care of patients with liver disease during the COVID-19 pandemic: EASL-ESCMID position paper. JHEP Reports, 2020: p. 100113.

17. Sterling, R.K., et al., Development of a simple noninvasive index to predict significant fibrosis in patients with HIV/HCV coinfection. Hepatology, 2006. 43(6): p. 1317–1325.

18. Vallet - Pichard, A., V. Mallet, and S. Pol, FIB - 4: A simple, inexpensive and accurate marker of fibrosis in HCV - infected patients. Hepatology, 2006. 44(3): p. 769–769.

19. Vallet - Pichard, A., et al., FIB - 4: An inexpensive and accurate marker of fibrosis in HCV infection, comparison with liver biopsy and fibrotest. Hepatology, 2007. 46(1): p. 32–36.

20. Kim, B.K., et al., Validation of FIB - 4 and comparison with other simple noninvasive indices for predicting liver fibrosis and cirrhosis in hepatitis B virus - infected patients. Liver International, 2010. 30(4): p. 546–553.

21. Li, Y., Y. Chen, and Y. Zhao, The diagnostic value of the FIB-4 index for staging hepatitis B-related fibrosis: a meta-analysis. PloS one, 2014. 9(8).

22. Nguyen, T.A., J.P. DeShazo, and A.J. Sanyal, Su1798 A High FIB-4 Score Predicts the Likelihood of Liver Related Outcomes in the General Population. Gastroenterology, 2013. 144(5): p. S-1006.

23. Davyduke, T., et al., Impact of Implementing a “FIB - 4 First” Strategy on a Pathway for Patients With NAFLD Referred From Primary Care. Hepatology communications, 2019. 3(10): p. 1322–1333.

24. Blackard, J.T., et al., HIV mono-infection is associated with FIB-4-a noninvasive index of liver fibrosis-in women. Clinical Infectious Diseases, 2011. 52(5): p. 674–680.

25. Kwo, P.Y., S.M. Cohen, and J.K. Lim, ACG clinical guideline: evaluation of abnormal liver chemistries. American Journal of Gastroenterology, 2017. 112(1): p. 18–35.

26. Tueller, S.J., R.A. Van Dorn, and G.V. Bobashev, Visualization of categorical longitudinal and times series data. Methods report (RTI Press), 2016. 2016.

27. Swihart, B.J., et al., Lasagna plots: a saucy alternative to spaghetti plots. Epidemiology (Cambridge, Mass.), 2010. 21(5): p. 621.

28. Liang, X., et al., Reprint of: Epidemiological serosurvey of Hepatitis B in China—Declining HBV prevalence due to Hepatitis B vaccination. Vaccine, 2013. 31: p. J21–J28.

29. Xia, G.-L., et al., Prevalence of hepatitis B and C virus infections in the general Chinese population. Results from a nationwide cross-sectional seroepidemiologic study of hepatitis A B, C, D, and E virus infections in China, 1992. International Hepatology Communications, 1996. 5(1): p. 62–73.

30. Docherty, A.B., et al., Features of 20133 UK patients in hospital with covid-19 using the ISARIC WHO Clinical Characterisation Protocol: prospective observational cohort study, bmj, 2020. 369.

31. Singh, S. and A. Khan, Clinical Characteristics and Outcomes of COVID-19 Among Patients with Pre-Existing Liver Disease in United States: A Multi-Center Research Network Study. Gastroenterology, 2020.

32. Ji, D., et al., Implication of non-alcoholic fatty liver diseases (NAFLD) in patients with COVID-19: a preliminary analysis. J Hepatol, 2020.

33. Gacouin, A., et al., Liver cirrhosis is independently associated with 90-day mortality in ARDS patients. Shock: Injury, Inflammation, and Sepsis: Laboratory and Clinical Approaches, 2016. 45(1): p. 16–21.

34. Donahoe, M., Acute respiratory distress syndrome: A clinical review. Pulmonary Circulation, 2011. 1(2): p. 192–211.

35. Ware, L.B. and M.A. Matthay, The acute respiratory distress syndrome. New England Journal of Medicine, 2000. 342(18): p. 1334–1349.

36. Emr, B., et al., Removal of inflammatory ascites is associated with dynamic modification of local and systemic inflammation along with prevention of acute lung injury: in vivo and in silico studies. Shock (Augusta, Ga.), 2014. 41(4): p. 317.

37. Arvaniti, V., et al., Infections in patients with cirrhosis increase mortality four-fold and should be used in determining prognosis. Gastroenterology, 2010. 139(4): p. 1246–1256. e5.

